# Effect of Adjunctive Intraoral Balance Appliance in the Usual Care of Patients with Chronic Temporomandibular Joint Disorders using Korean Medicine: Study Protocol for a Randomized Controlled Trial

**DOI:** 10.1101/2024.06.18.24309139

**Authors:** Woo-Chul Shin, Se yun Kim, Whisung Cho, Jaehyun Park, Hyungsuk Kim, Won-Seok Chung, Mi-Yeon Song, Jae-Heung Cho

## Abstract

**Introduction:** Temporomandibular disorders (TMD) are a group of conditions that affect the temporomandibular joint, masticatory muscles, and associated structures, often leading to pain, dysfunction, and a significant impact on quality of life. Epidemiological studies have estimated that up to 75% of the population in the United States exhibit at least one sign of TMD. Although conservative treatments such as acupuncture and occlusal splints have been recommended, evidence for their effectiveness remains inconclusive, and the combined effects of these interventions are not well understood.

This study aims to compare the efficacy of an intraoral balance appliance (IBA) combined with standard Korean medicine care versus Korean medicine care alone in patients with chronic, painful TMD with myalgia.

**Materials and Methods:** A single-center, two-arm, parallel, evaluator-blinded, randomized controlled trial (RCT) with a 1:1 allocation ratio will be designed to test the interventions. Seventy-six TMD patients with myalgia will be recruited and randomized. The Interventions will include manual acupuncture and physical therapy in both groups, with the addition of IBA in the treatment group.

**Discussion:** The outcomes will be measured using various scales such as the numeric rating scale for pain and bothersomeness, jaw functional limitation scale, and quality of life indicators.

**Conclusion:** The trial is expected to provide evidence of the efficacy of combining the usual Korean medicine care and IBA in managing chronic TMD myalgia. Despite certain limitations such as the short intervention period and lack of standardized splint therapy, this RCT will contribute valuable data to guide the future treatment of TMD with myalgia.

**Trial registration:** Clinical Research information Service (CRiS) ID: KCT0008906. Registered on October 30, 2023

## Introduction

The temporomandibular joint (TMJ) is the only bilateral synovial joint in the human body [1]. This allows opening and closing of the mouth for mastication, which is essential for the life-sustaining functions of the body. Temporomandibular disorders (TMD) are a subgroup of craniofacial pain problems that involve the TMJ, masticatory muscles, and associated head and neck musculoskeletal structures [2]. Temporomandibular disorders can lead to a cluster of symptoms, including jaw pain, facial pain, headache, and associated psychological and psychosocial problems. Given the varied subgroups of symptoms and signs present in TMD, the etiological factors are recognized as multifactorial, encompassing biological, behavioral, environmental, social, emotional, and cognitive influences, either independently or in combination, contributing to the disorder. According to the diagnostic criteria for TMD (DC/TMD), TMD can be classified as either intra- or extra-articular [3]. Extra-articular conditions, which usually involve musculature problems surrounding the TMJ, are the most common causes of TMD, accounting for at least 50% of TMD cases [4]. Additionally, among the various signs and symptoms of TMD, pain-related TMD is more common and significantly affects an individual’s daily activities. Epidemiological studies have shown that 40–75% of the population exhibits at least one of the signs of TMD [2]. Additionally, the OPPERA project (Orofacial Pain: Prospective Evaluation and Risk Assessment) estimated the annual incidence of TMD to be approximately 4% in the US population [5], with a 20.4% prevalence of chronic pain, indicating that TMD represents a major public health burden.

One suggested initial approach for TMD, particularly of myogenous origin, is conservative management because of its multifaceted causes and unknown precise pathogenesis. Conservative treatment modalities include counseling, medication, exercise, occlusal splints, and acupuncture [4]. Among the treatment modalities, acupuncture has garnered widespread adoption, particularly in East Asian countries, owing to its relatively few adverse effects and notable analgesic efficacy. Multiple controlled trials have reported the efficacy of acupuncture on pain conditions, such as myofascial pain, tension headache, and fibromyalgia. [6] Similar to myofascial pain conditions, TMD with myalgia is often associated with tender points in related masticatory muscles. Systematic reviews have shown that acupuncture reduces the signs and symptoms of myofascial TMD pain; however, the quality of the included studies was low [7, 8]. Another conservative treatment option is occlusal splints. One of the main pathologies of TMD is mechanical displacement, usually within the disc-condyle relationship. Various splints are used to restore static and dynamic occlusions, relax the relevant musculature, and reduce physiological stress in joint structures [9]. Several systematic reviews have indicated the benefits of occlusal splints in treating TMD [10]. Moreover, a recent network meta-analysis demonstrated that occlusal splints are one of the most effective treatment modalities for TMD, yielding significant benefits in both short and intermediate terms [11].

Despite the reported advantages of various treatments for TMD, a definitive consensus regarding the most efficacious treatment is still lacking as insufficient conclusive evidence is unavailable. Consequently, a multidisciplinary approach is usually recommended in clinical practice [12]; however, many interventions have not yet been rigorously tested.

We aim to compare the efficacy of incorporating an intraoral balance appliance (IBA) alongside the usual care for patients with chronic painful TMD using Korean medicine and evaluate its effectiveness on patient’s functional, clinical outcomes (pain, bothersomeness, vertical range of motion (ROM), graded chronic pain scale (GCPS), jaw functional limitation scale (JFLS), patient health questionnaire-9 (PHQ-9), quality of life (EQ-5D-5L), and patient global impression of change (PGIC)), and adverse events (AEs).

## Materials and methods

### Trial design

This is a single-center, two-arm, parallel, evaluator-blind randomized controlled trial with a 1:1 allocation ratio. The study will be performed at Kyung Hee University Medical Center, Korean Medicine Hospital, Korea. The reporting of the protocol will adhere to the Standard Protocol Items: Recommendations for Interventional Trials (SPIRIT) (S1 table) [13]. The trial follows the Consolidated Standards of Reporting Trials (CONSORT) reporting guidelines and the Standards for Reporting Interventions in Clinical Trials of Acupuncture (STRICTA) recommendations (S2 table) [14].

### Ethics approval

Ethical approval was obtained from the Institutional Review Board of Kyung Hee University Korean Medicine Hospital Institutional Review Board (IRB number: KOMCIRB 2023-06-001). We will obtain informed consent from each eligible participant, and collect data anonymously to maintain privacy. This study will be conducted in full conformity with the 1964 Declaration of Helsinki and all subsequent revisions and in accordance with the Good Clinical Practice Guidelines. The protocol will be registered at the Clinical Research information Service (CRiS), Republic of Korea, on October 30, 2023 (CRiS number KCT0008906) before enrollment of the first participant.

### Sample size calculation

This study is a prospective study that will compare the effects of different IBA. Referring to a previous study that compared treatments for painful masticatory muscles [15], an appropriately powered full-scale sample size will be estimated using the mean difference of the numeric rating scale (NRS) for pain owing to TMD with myalgia between groups at the primary endpoint. The effect size *d* according to the change in the reference study is 0.74, and with 80% power, 0.05 alpha, and a two-sided test, the number of samples required for each group will be 30. The total sample size required, when considering a dropout rate of 20%, will be 76 participants, with 38 participants in each group. The sample size will be determined using G*Power 3.1.7 software.

### Methods of recruitment

Recruitment will be accomplished by issuing press releases about the study, advertisements in free newspapers, and displaying posters within and outside Kyung Hee University Medical Center Korean Medicine Hospital in Seoul, Korea. The estimated recruitment period is 12 months.

### Eligibility criteria

The inclusion criteria will be as follows:

1. Participants aged between 19–70 years on the date of signing the informed consent form.
2. Participants with unilateral or bilateral TMJ pain.
3. Participants with a NRS of ≥4 at the site of TMJ pain (based on the side with more pain for participants with bilateral pain).
4. Participants with persistent or intermittent TMJ pain for ≥3 months.
5. Participants with a diagnosis of TMD with myalgia according to the DC/TMD diagnostic criteria.
6. Participants who voluntarily agree to participate in the study and sign a written informed consent form prior to any study-related procedures.

The exclusion criteria will be as follows:

1. Current pain episode caused or exacerbated by traumatic injuries such as motor vehicle accidents.
2. Individuals who do not meet the diagnosis of TMD with myalgia according to the DC/TMD diagnostic criteria.
3. Undergone surgery related to TMJ.
4. Concomitant medical conditions that may interfere with the efficacy of treatment or interpretation of results (e.g., rheumatoid arthritis, tumor, stroke, and myocardial infarction)
5. Participants currently taking steroidal agents, immunosuppressants, psychiatric medications, and other medications that may affect the results of the trial.
6. Participants who have taken medications that may affect pain, such as non-steroidal anti-inflammatory drugs within the past week, or have applied orthodontic appliances in the past two weeks, regardless of the condition.
7. Participants who are pregnant, planning to become pregnant, or breastfeeding women.
8. Participated in another clinical trial within one month before the date of enrollment, or plans to participate in another clinical trial during the study participation and follow-up period.
9. Other reasons deemed by the investigator that may present difficulties to participate in the trial.

Participants in the trial will voluntarily sign the written informed consent forms in accordance with the Declaration of Helsinki and Guidelines for Good Clinical Practice. The consent form will be collected by the participating Korean medicine doctors (KMDs) or a participating clinical research coordinator.

### Randomization and allocation strategy

Randomization will be conducted using computer-generated random numbers. A statistician who is not involved in the intervention process will generate computer-generated random numbers. Blocked randomization will be used to ensure a balance between the two groups, with the block size kept confidential from those involved in patient enrollment or intervention assignment. The randomized numbers will be assigned to screen the participants in the order of recruitment. The designated investigator will provide the intervention arm to the practitioners. The generated random numbers will be sealed in an envelope and stored by an investigator who will not engage with the participants.

### Dropout and withdrawal

If any participants voluntarily withdraw their consent at any point during the study or if an adverse event (AE) that is considered harmful to the participants is observed, the investigator will temporarily or permanently discontinue the study for the respective participants. Detailed reasons for any interrupted study trials will be recorded by the investigator. If early withdrawal owing to an AE occurs, appropriate treatment will be provided and the investigator will continue to monitor the participant until the AE resolves. In case of consent withdrawal, no further follow-up will be conducted. The investigator may also terminate the study if the participant receives a procedure, medication, or other treatments that could affect the study without permission. Study participants who withdraw before treatment initiation will be considered screening failures. The investigators will make every effort to obtain information about the patients who drop out and record the reason for discontinuation in the electronic case report forms (eCRF).

### Blinding

This will be an open-label study with participants and practitioners being aware of treatment allocation. Only the outcome evaluator will be blinded to the intervention arm. Therefore, unblinding will be unnecessary in any circumstances.

### Participants’ timeline

The duration of the study will be four weeks. A timeline of this study is shown in Fig 1.

**Fig 1.**
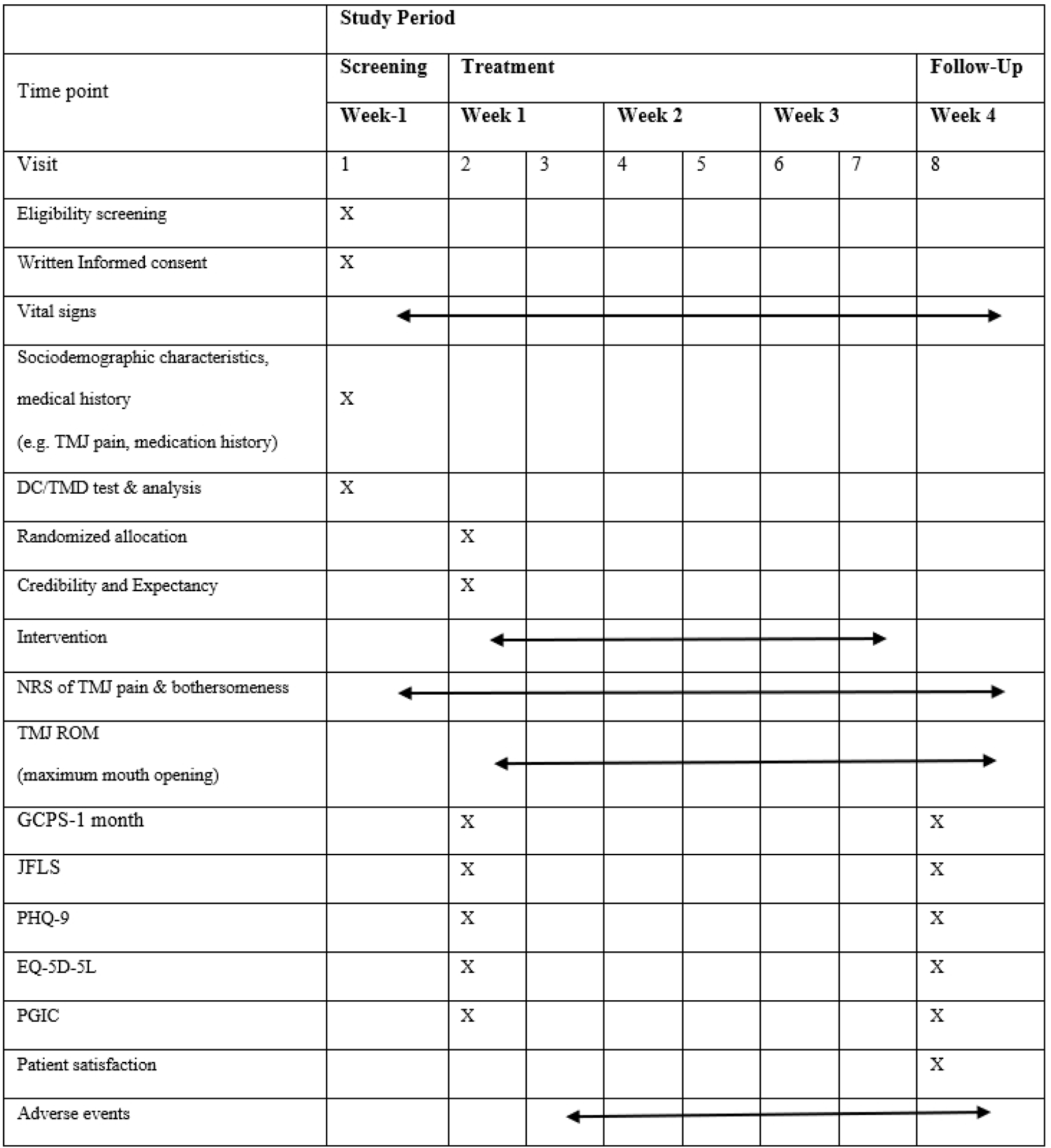
SPIRIT schedule of enrollment, interventions, and assessments.

### Interventions

Participants will be randomized into treatment and control groups. Both groups will receive the usual care of Korean medicine consisting of manual acupuncture and physical therapy. The treatment group will receive additional IBA (JinBiotech®, Cheonan, Korea). Owing to the inherent nature of splint therapy, creating a placebo group is fundamentally impractical; therefore, it will not be included in this study. Acupuncture and physical therapy will be administered twice a week for three weeks, with a total of six treatment sessions for both the treatment and control groups.

Manual acupuncture will be performed at four essential acupuncture points: Xiaguan (ST7), Jiache (ST6), Ermen (TE21), and Yifeng (TE17). Additional ‘ashi points,’ which are tender points in associated musculature, will be determined by the participating KMDs. Manual acupuncture will be administered using disposable sterile stainless-steel needles (40 × 0.25 mm; Dong-bang Medical, Seongnam, Korea) after skin sterilization with the participants lying face down. The needles will be inserted perpendicularly or obliquely, depending on the acupuncture point, to a depth of 5–30 mm, and left in place for 15 min. More detailed procedures based on STRICTA are attached in S2 Table. Infrared radiation (IR 3000; Haedong Medical, Ulsan, Korea) will be simultaneously applied to the area where acupuncture treatment has been performed for 15 min.

Physical therapy will be performed using an interferential current therapy (ICT) device (Nihon Medix Model 5402, Japan). The participants will be asked to lie down in the prone position with the cervical area unclothed. Four self-adhesive 6×6 cm electrodes will be placed using a crossed pattern in the lower cervical area. The intensity will be varied according to the individual tolerance. The ICT stimulation will be applied for 15 min per session. The participants will be instructed to immediately report any discomfort during treatment.

The participants in the treatment group will be instructed to apply IBA for a minimum of 6 h a day over three weeks. The IBA will be positioned in the participant’s mouth with the tongue tag portion facing upward, and the outer centerline of the appliance will be aligned with the middle of the upper teeth. The participants will be instructed to gently place their teeth on the occlusal plane without clenching or applying force to their molars while wearing the device. A GMP certification (KTR-AABB-16978) has been obtained for the device from the Korea Testing & Research Institute.

During the treatment period (Weeks 1–3), any procedure or medication aimed directly at relieving painful TMD will be prohibited. However, ibuprofen will be allowed as a relief medication during the study for all participants, and the dosage will be self-reported. Concomitant care will be allowed for the subsequent follow-up period (week four). Any treatment administered during the follow-up period will be documented during the last visit.

### Practitioner background

Acupuncture treatments will be conducted by KMDs licensed by the Korean Ministry of Health and Welfare, who specialize in rehabilitation medicine of Korean medicine and have at least five years of clinical experience. Techniques for the entire treatment procedure will be standardized by practitioners.

### Measurements

At screening, the assessment will include demographics (age and sex), anamnesis (underlying illness and medical and treatment histories), physical examination (muscle palpation), and ROM of the TMJ. The database will include the following information after enrollment and allocation:

1. Patient characteristics: height (cm), weight (kg), and vital signs (body temperature, blood pressure, and pulse rate).
2. Outcomes: NRS for pain and bothersomeness, vertical ROM of the TMJ, GCPS, JFLS, PHQ-9, EQ-5D-5L, PGIC, patient satisfaction, credibility and expectancy, and AE.

The primary outcome of the study will be the NRS score for pain and bothersomeness assessed at the final visit at week four. Participants will select a number between 0 and 10 that best represents their current pain intensity, with 0 indicating no pain and 10 denoting the most severe pain imaginable. In cases where participants complain of pain in both TMJs, the side with the most severe pain will serve as the reference.

The ROM of the TMJ is the vertical ROM measured with a ruler as the maximum range of voluntary opening without pain. The test will be performed in accordance with the clinical examination protocol provided by the International DC/TMD consortium [16].

The GCPS-1-month version uses the same item structure as the original GCPS-6-month scale, published in 1992. The DC/TMD includes the GCPS-1-month version because of its recognized clinical utility in assessing pain persistence. The scale has been evaluated for reliability and validity, and it consists of seven items that assess pain intensity and pain-related disabilities over the past 30 days [3].

The JFLS is a global measure of functional limitation derived from the Research Diagnostic Criteria for TMD Checklist and the Mandibular Functional Impairment Questionnaire. This scale comprises 20 items that assess limitations in mastication, jaw mobility, and verbal and emotional expressions. Previous studies have reported its reliability and validity [17].

The PHQ-9 has the advantage of being aligned with the diagnostic criteria for depression in the fourth edition of the Diagnostic and Statistical Manual of Mental Disorders with good sensitivity and specificity. The measurement is short and practical, with only nine items. The reliability and validity of the Korean version of the PHQ-9 have been validated in numerous studies [18].

The EQ-5D-5L, developed in 2010, is a five-level version of the EQ-5D and is a broadly used generic multi-attribute health utility instrument. Composed of five items, the EQ-5D-5L is currently available in >130 languages, and it demonstrated improved psychometric properties over the EQ-5D-3L version. [19]

The PGIC and patient satisfaction questionnaires will be used to assess the subjective responses of patients to symptom changes after treatment. The PGIC is a seven-point transition scale used to evaluate patients’ perceptions of their overall change in symptoms after intervention [20]. The patient satisfaction questionnaire consists of three questions, each rated on a nine-point Likert scale. Additionally, the nine-point Likert scale will be used to assess the participants’ credibility and expectancy toward the treatment.

### Data collection and management

Electronic case report forms will be developed and validated to collect all study-related information. The designated site staff will document participant data in a local participant file. The investigators will manage and conduct quality control of the data according to their standard operating procedures. The participant files will serve as source data for the study, and the data will be entered into the eCRF. The eCRF platform will be provided by the National Clearinghouse for Korean Medicine (NCKM). To ensure the integrity of the study data, only one individual will be allowed to assess the eCRF at a time, and both the eCRF and supporting documents will be subjected to cross-checking. Monitoring will be scheduled three times during the study: initial monitoring upon the selection of the initial patient, interim monitoring at any point during the study duration, and final monitoring after the study. Monitoring will be conducted by personnel independent of the study process. All documents related to the study will be kept in a designated secured storage inside the study management department. All records will be retained for three years after the end of the clinical study. After the retention period, the records will be terminated to ensure confidentiality.

### Statistical methods

Sociodemographic characteristics, treatment credibility, and expectations will be assessed for each group. Primary analysis will involve an intention-to-treat approach, while per-protocol analysis will be conducted simultaneously if necessary. To compare the differences between the two groups, Student’s t-test will be used for continuous variables, and the chi-square test or Fisher’s exact test will be performed for categorical variables. The efficacy variables will be the difference in the change in continuous outcomes between the two groups from baseline to each evaluation time point. A repeated-measures analysis of variance (RM-ANOVA) or a mixed model for repeated data will be performed to test for differences in trend changes across visits. Additionally, an analysis of covariance (ANCOVA) will be performed using the baseline values of each variable and covariate factors that are statistically different between the groups at baseline, with the group as a fixed factor. The analysis will examine the proportion of patients whose pain NRS scores decrease by half or more from baseline at each evaluation time point. A Kaplan–Meier survival analysis will assess the duration from randomization to achieving a reduction in NRS pain scores by less than half, and the resulting curves will be compared using the log-rank test. A Cox model will be employed to compare the rate of TMJ pain reduction to less than half by calculating the hazard ratio. Any missing data from participants who drop out will be handled mainly by multiple imputations, and the last observation carried forward will be used for the sensitivity analysis.

The significance level for all the analyses will be set at 0.05. All the statistical analyses will be performed using the SAS version 9.4 statistical package (SAS Institute, Cary, NC, USA). Subgroup analyses will be conducted, if necessary. No interim analyses will be planned for the study as the sample size calculation will be tailored for the treatment period, which will make it improbable to achieve a statistically significant difference between the groups before the primary endpoint.

### Confidentiality

Investigators with direct access to the source data will take all necessary precautions to ensure the confidentiality of the information, and participant confidentiality will be strictly maintained as far as possible under the law and hospital policy. Each participant that will be enrolled in the study will be encrypted by assigning a unique identification number to protect the confidentiality of personal information. The medical information of participants obtained in the study will be confidential and may be disclosed to third parties, as permitted by the patient’s signed consent form. Additionally, medical information may be provided for treatment purposes to the appropriate healthcare personnel responsible for patient welfare. To the extent necessary, data from the study may be presented upon request by monitoring personnel or by the Institutional Review Board (IRB) of the clinical research organization.

### Adverse event reporting and harms

In a clinical study, an AE is denoted as an undesirable and unintended sign, symptom, or disease that occurs after a procedure and is not necessarily causally related to the procedure. Serious AE (SAE) are as follows:

1. Causes death or is life-threatening.
2. Requires hospitalization or extended hospitalization.
3. Causes persistent or significant disability or reduced functioning.
4. Other medically significant circumstances.

Phenomena expected to be AE will be fully recorded in detail (signs, symptoms, onset date, duration, etc.), and evaluation of the AE will be recorded in a separate case report form by the investigator. The causal relationship with the intervention will be evaluated according to the World Health Organization-Uppsala Monitoring Centre causality categories [21]. The degree of symptoms will be evaluated using the three-level classification method of Spilker [22].

### Protocol amendments

If changes to the study protocol are required, all changes will be approved by the IRB of the clinical research organization before implementation, except for those necessary to eliminate immediate participant risk and administrative changes.

### Trial status and dissemination plans

Currently, the study protocol, eCRF, and relevant documents are being reviewed before trial initiation. The protocol is version 2.3 (2023-08-04). The estimated date of recruitment completion is December 2024.

The results of this study will be actively disseminated through manuscript publications and conference presentations. Regardless of whether the study is completed or prematurely terminated, the clinical study report will be provided to the IRB of the clinical research organization.

## Discussion

In 1934, Dr. James Costen published a paper describing a syndrome characterized by ear pain, tinnitus, vertigo, and limited jaw movement, which he attributed to abnormalities in the TMJ. Despite differences in modern terminology, this syndrome is considered one of the earliest descriptions of what we now recognize as TMD [23]. Almost a century has passed, yet the prevalence of TMD has remained fairly consistent or has even increased over the years with no definitive treatment, posing a significant burden both individually and globally [24].

Despite the promising efficacy reported by several systematic reviews of various conservative treatments for TMD, such as acupuncture, occlusal splinting, photomodulation, laser, biofeedback, cognitive behavioral training, hypnosis, botulinum toxin A, exercise, and medication, many studies have reported conflicting results with insufficient evidence [25]. However, recent guidelines for the management of TMD recommend acupuncture and splint therapy because conservative approaches should be pursued first [26]. Clinically, acupuncture is used in various forms including traditional acupuncture, dry needling, electroacupuncture, warm-needle acupuncture, and laser acupuncture. Regardless of the study type, recent systematic reviews have revealed that acupuncture, whether used independently or as an adjunct to other treatments, can significantly improve the effect rate and pain intensity in patients with TMD compared with inactive controls, although the quality of the evidence was low [7, 27]. A systemic review comparing conservative therapeutic modalities for TMD concluded that occlusal splints, alone or in combination with other interventions, had a positive effect on short-term TMD pain reduction [25]. However, owing to the inherent heterogeneity of splint studies, such as the various types of splints used, delivery methods, and study designs, it is challenging to draw firm conclusions [28]. Few studies have assessed the efficacy of combining acupuncture with splint therapy for the treatment of TMD. One study with a design similar to ours compared acupuncture versus acupuncture plus splint therapy for painful TMD over four weeks [29]. Although the combination of interventions did not show significant differences between the groups, the splint group demonstrated an accelerated decrease in pain and improved short-term efficacy.

The potential limitations of the study are as follows: 1) the intervention period was short, with no long-term follow-up evaluation scheduled, which means that only short-term effects can be determined; 2) since there is no standardized splint therapy for the treatment of painful TMD, the results of this study will be confined to a specific splint, the IBA, and cannot be extrapolated to other types of splints. Nevertheless, this randomized, assessor-blind, controlled trial will provide high-quality data on the efficacy of combined treatment with acupuncture and the IBA for TMD with myalgia and may help to guide future directions for the treatment of TMD with myalgia.

## Abbreviations

AE: Adverse event

ANCOVA: Analysis of covariance

CONSORT: Consolidated standards of reporting trials

CRiS: Clinical Research information Service

DC/TMD: Diagnostic criteria for temporomandibular disorders

eCRF: Electronic case report form

EQ-5D-5L: 5-Level EuroQol-5 dimension

GCPS: Graded chronic pain scale

IBA: Intraoral balance appliance

IRB: Institutional review board

JFLS: Jaw functional limitation scale

KMD: Korean medicine doctor

NCKM: National Clearinghouse for Korean Medicine

NRS: Numeric rating scale

PHQ-9: Patient health questionnaire-9

PGIC: Patient global impression of change

SAE: Severe adverse event

STRICTA: Standards for reporting interventions in clinical trials of acupuncture

TMD: Temporomandibular joint disorder

TMJ: Temporomandibular joint

RM-ANOVA: Repeated measure analysis of variance

ROM: Range of motion

## Author’s contributions

All the authors have read and approved the final manuscript. Conceptualization: WCS, SYK, and JHC; methodology: WCS, SYK, and JHC; writing-original draft preparation: WCS and JHC; writing-review and editing: WCS, WC, JP, HK, WSC, MYS, and JHC; visualization and project administration: JHC.

## Data Availability

No datasets were generated or analysed during the current study. All relevant data from this study will be made available upon study completion.

